# Ketamine improves anhedonic phenotypes across species: Translational evidence from the Probabilistic Reward Task

**DOI:** 10.1101/2025.06.02.25328447

**Authors:** Mario Bogdanov, Jason N. Scott, Shiba M. Esfand, Brian W. Boyle, Ty Lees, Mohan Li, Sarah E. Woronko, Samantha R. Linton, Amaya R. Jenkins, Courtney Miller, Shuang Li, Paula Bolton, Daniela B. Radl, Thomas J. Kornecook, Robert C. Meisner, Brian D. Kangas, Diego A. Pizzagalli

**Affiliations:** Center for Depression, Anxiety, and Stress Research, McLean Hospital, Belmont, MA, USA; Department of Psychiatry, Harvard Medical School, Boston, MA, USA; Psychiatric Neurotherapeutics Program, McLean Hospital, Belmont, MA, USA; Behavioral Biology Program, McLean Hospital, Belmont, MA, USA; Neurocrine Biosciences Inc., San Diego, CA, USA; Acute Psychiatric Service, Massachusetts General Hospital, Boston, MA, USA; Noel Drury, M.D. Institute for Translational Depression Discoveries, University of California, Irvine, CA, USA

**Keywords:** reward responsiveness, ketamine, anhedonia, treatment-resistant depression, rats, humans

## Abstract

**Background:** Ketamine is increasingly used as a therapeutic option for treatment-resistant depression (TRD) due to its rapid antidepressant properties, yet the mechanisms underlying these effects remain elusive. Preclinical evidence suggests ketamine acts on neural pathways implicated in reward processing, but translational efforts have proven challenging, due to a lack of paradigms allowing for analogous assessment of depressive phenotypes across species. Here, we investigated the effects of a single, subanesthetic dose of ketamine on reward responsiveness in individuals with TRD and chronically-stressed rats using functionally identical tasks.

**Methods:** Human participants completed the Probabilistic Reward Task (PRT) twice within 48h, either without intervention (healthy controls, n=36, 26 women) or 24h before and after ketamine administration (individuals with TRD, n=24, 16 women). Rats (all male) completed a reverse-translated version of the PRT on three separate days (healthy controls, n=10) or before and after chronic stress exposure as well as 2h and 24h after ketamine administration (experimental group, n=10).

**Results:** Ketamine significantly increased response bias toward the more frequently rewarded stimulus in both humans and rats, resulting in levels comparable to healthy controls 24h post-administration. Exploratory analyses in humans suggested that this effect was strongest among more anhedonic individuals. Furthermore, in both species, ketamine had no effect on measures of overall task performance, suggesting ketamine selectively affected reward learning rather than general cognition.

**Conclusions:** Our results indicate a shared behavioral mechanism through which ketamine alleviates anhedonic behaviors and offers important implications for the treatment of people suffering from anhedonia in TRD and related psychopathologies.

## Background

Despite continued research on the neurobiology and treatment of Major Depressive Disorder (MDD), approximately 30% of depressed individuals do not benefit from multiple conventional treatment options within a given depressive episode and thus are considered to have treatment-resistant depression (TRD) (1,2). TRD is associated with increased rates of hospitalization and suicidality as well as an overall reduced quality of life, emphasizing the need to identify effective interventions (3). Following reports of rapid and robust antidepressant effects of ketamine, a non-competitive N-methyl-D-aspartate (NMDA) receptor antagonist (4), recent work has evaluated the therapeutic use of ketamine in people with TRD (5–9). Yet, while the acute clinical effects of ketamine are well-characterized, its effects on cognitive, behavioral, or affective mechanisms that may modulate its antidepressive response remain poorly understood.

Ketamine has been proposed to be particularly effective for alleviating anhedonia - defined as the lack of interest in or pleasure from previously enjoyable activities and one of the cardinal symptoms of MDD (5,10–16). Anhedonia is associated with altered reward processing, including reduced responsiveness to anticipated or received rewards and impaired learning from rewarding outcomes (17,18). Critically, symptoms of anhedonia are generally not improved by common first-choice pharmacological treatments, such as Selective Serotonin Reuptake Inhibitors (SSRIs; (19)). In contrast, ketamine, administered either by injection of racemic (*R,S*) ketamine or intranasally as esketamine (the (*S*)-enantiomer of ketamine), has been shown to reduce self-reported and clinician-assessed anhedonia within hours (6,14,20,21). Moreover, preclinical work and neuroimaging studies have provided ample evidence for rapid and widespread effects of ketamine on brain structures associated with reward processing, including the anterior cingulate cortex (ACC), ventromedial prefrontal cortex, orbitofrontal cortex, nucleus accumbens, and (lateral) habenula, as well as changes in activity, connectivity, and synchronicity of large-scale brain networks (6,22–31). These changes are thought to be mediated by molecular mechanisms that, ultimately, result in increased dopaminergic tone in mesocorticolimbic pathways critical for reward learning and incentive motivation (8,32–34).

The current understanding of the mechanisms underlying antidepressant effects of ketamine stems primarily from preclinical models, and it is unclear whether these findings apply to humans. A major challenge in translational efforts is the lack of behavioral endpoints that are functionally analogous across species (35,36). For example, human studies often use changes in self-reported symptom severity to indicate treatment success (5,6,20). Conversely, common experimental paradigms probing anhedonic-like phenotypes in animals, (e.g., sucrose preference task and intracranial self-stimulation) cannot easily be employed in humans (37).

To address this issue, we recently reverse-translated the Probabilistic Reward Task (PRT), an established paradigm to investigate reward learning in humans (38) and a recommended task within the Positive Valence Systems in the RDoC framework (39), for rodents and nonhuman primates (40–42). In the PRT, subjects make relatively difficult visual discriminations between two stimuli. Importantly, unbeknownst to subjects, correct responses to one stimulus are rewarded three times more frequently (rich stimulus) than correct responses to the other stimulus (lean stimulus). Among healthy humans and experimental animals, this asymmetric reinforcement schedule reliably induces a response bias toward the rich stimulus (43). Conversely, humans with anhedonia and animals exposed to early-life adversity or chronic stress display blunted response bias (44–48).

Leveraging the PRT, the current study aimed to investigate whether a single, subanesthetic dose of ketamine would enhance blunted reward responsiveness in individuals with TRD and rats with anhedonic phenotypes. Therefore, rats were exposed to a validated chronic stress paradigm known to induce anhedonic-like behavior in the PRT (48), before receiving a single dose of ketamine. We compared their PRT performance before and after stress exposure as well as after ketamine administration to that of unstressed control rats. Similarly, we tested response bias in treatment-seeking individuals with TRD 24 hours before and 24 hours after their first administration of ketamine and compared their performance to healthy controls.

Based on prior findings (15,16,20,49), we hypothesized that ketamine would lead to a rapid prohedonic effect (i.e., significantly increased response bias) in both species. In rats, we expected that ketamine would rescue the experimentally induced reduction in response bias in the chronic stress group, with a return to levels observed at the pre-stress baseline and comparable to those in non-stressed controls. For TRD participants, we expected a decrease in self-reported depressive symptom severity following ketamine treatment. Furthermore, we expected a lower response bias in TRD participants during their first session (i.e., pre-treatment) compared with healthy controls and an increase in response bias in their second session (i.e., post-treatment) to levels comparable to those of healthy controls. Finally, we explored whether treatment-related changes in response bias correlated with individual differences in self-reported anhedonia.

## Methods and Materials

### Animals

Twenty adult male Long-Evans rats were obtained from Charles River Laboratories (Wilmington, MA). To establish sweetened condensed milk as a reinforcer, rats were restricted to approximately 10-15 grams of rodent chow given daily after the experimental session. Water was available ad libitum in the home cage. The study protocol was approved by the Institutional Animal Care and Use Committee at McLean Hospital in accordance with established guidelines (50).

### Human Participants

As part of a larger study, we recruited 24 individuals with TRD (16 females, 8 males, *M* age ± *SD* = 44.35 ± 15.86 years, range = 21 – 69) through McLean Hospital’s ketamine service and 36 psychologically healthy controls (26 female, 10 male, *M* age ± *SD* = 33.18 ± 14.49 years, range = 19 – 68) from the Greater Boston area. All participants were ketamine-naïve and screened for depressive symptoms prior to study begin using the Mini International Neuropsychiatric Interview (MINI) (51) and the Hamilton Depressive Rating Scale (HAMD) (52). Participants also completed the Beck Depression Inventory (BDI-II) (53), Quick-Inventory of Depression (QIDS) (54) and Snaith-Hamilton Pleasure Scale (SHAPS) (55). To ensure PRT data quality, well-established quality checks were applied (56,57), resulting in the exclusion of 8 healthy control and 6 TRD participants for a final sample size of n = 46 (28 HC, 18 TRD). Demographic and clinical characteristics are summarized in Table 1 (see also *Supplement*).

**Table 1.** Demographic and clinical characteristics (human samples)

| | HC<br>(n = 28) | TRD<br>(n = 18) | *t* / $\chi^2$ (df) | *p* | |
| --- | --- | --- | --- | --- | --- |
| Screening session |  |  |  |  |  |
| % female (n) | 78.6 (22) | 66.7 (12) | 0.31 (1) | .580 |  |
| Age (years, mean ± SD) | 33.49 ± 14.36 | 45.42 ± 16.62 | -2.50 (32.52) | .018 | * |
| Education, (years, mean ± SD) | 16.29 ± 4.23 | 16.78 ± 3.56 | -0.42 (40.77) | .673 | |
| % White (n) | 64.3 (18) | 100 (18) | 6.90 (1) | .009 | ** |
| % Asian (n) | 35.7 (10) | – |  |  |  |
| HAMD (mean ± SD) | 0.21 ± 0.49 | 16.89 ± 4.86 | -14.09 (17.22) | <.001 | *** |
| QIDS (mean ± SD) | 0.11 ± 0.31 | 15.56 ± 3.29 | -19.43 (17.19) | <.001 | *** |
| BDI-II (mean ± SD) | 0.86 ± 1.43 | 35.78 ± 6.66 | -21.91 (18.02) | <.001 | *** |
| SHAPS (mean ± SD) | 19.89 ± 6.29 | 38.88 ± 5.29 | -10.63 (38.21) | <.001 | *** |
| First experimental session |  |  |  |  |  |
| HAMD (mean ± SD) | 0.33 ± 0.78 | 14.83 ± 4.27 | -14.24 (17.77) | <.001<sup>a</sup> | *** |
| QIDS (mean ± SD) | 0.15 ± 0.46 | 13.24 ± 3.73 | -14.38 (16.30) | <.001<sup>a</sup> | *** |
| BDI-II (mean ± SD) | 0.70 ± 1.14 | 33.47 ± 8.79 | -15.29 (16.34) | <.001<sup>a</sup> | *** |
| SHAPS (mean ± SD) | 18.86 ± 5.45 | 37.33 ± 6.22 | -10.31 (32.90) | <.001<sup>a</sup> | *** |
| Second experimental session |  |  |  |  |  |
| HAMD (mean ± SD) | 0.18 ± 0.61 | 10.24 ± 5.12 | -8.07 (16.28) | <.001<sup>a</sup> | *** |
| QIDS (mean ± SD) | 0.21 ± 0.63 | 8.59 ± 4.35 | -7.90 (16.41) | <.001<sup>a</sup> | *** |
| BDI-II (mean ± SD) | 0.62 ± 1.13 | 27.50 ± 9.85 | -11.53 (17.31) | <.001<sup>a</sup> | *** |
| SHAPS (mean ± SD) | 19.57 ± 5.95 | 35.06 ± 6.50 | -8.14 (34.03) | <.001<sup>a</sup> | *** |

**Note.** BDI-II = Beck Depression Inventory, HAMD = Hamilton Depression Rating Scale; HC = Healthy control; QIDS = Quick-Inventory of Depression; SHAPS = Snaith-Hamilton Pleasure Scale; TRD = Patient with Treatment Resistant Depression. <sup>a</sup> Bonferroni-corrected *p*-values for post-hoc *t*-tests following a two-way ANOVA (Group × session).

All participants provided written informed consent prior to participation and received monetary compensation of $75 for the screening visit, and either $75 or $125 per day for the experimental sessions (remuneration increased part-way through the study to aid recruitment). Study procedures were conducted in accordance with the Declaration of Helsinki and were approved by the Massachusetts General Brigham Healthcare Institutional Review Board.

### Drug

For the rat protocol, ketamine hydrochloride was obtained from Sigma-Aldrich (St. Louis, MO), dissolved in 0.9% saline solution and administered via subcutaneous injection in volumes of 0.5 mL or less 2h before the experimental session. The dose (10 mg/kg) was based on 1) previous studies indicating its approximation in rats with the clinically efficacious outcomes in humans (58–60), and 2) its production of peak prohedonic effects in previous rat PRT studies (49).

Human participants received a subanesthetic ketamine dose of 0.5mg/kg, delivered intravenously over 40 minutes. This procedure is in line with earlier human ketamine treatment studies (7,61) and followed current recommendations (62), highlighting this dose as being effective, safe, well-tolerated, and leading to no/minimal general anesthetic effects, respiratory problems, or cognitive impairments 24 hours post-administration (63,64). Injections were administered at McLean Hospital’s ketamine service as part of patients’ treatment plan.

### Probabilistic Reward Task (PRT)

Procedures for the human PRT have been described elsewhere (38,44) (Figure 1). Briefly, participants completed three blocks of 100 trials each in which they were presented with schematic face stimuli featuring a “long” (13.0 mm) or “short” (11.5 mm) mouth. Participants were asked to indicate the length of the mouth stimulus by button press as fast as possible. Importantly, the reward schedule in the PRT was asymmetric, so that correct responses to one stimulus (“rich stimulus”, 60%) were rewarded three times more frequently compared to the other (“lean stimulus”, 20%). Conditions (i.e., whether long or short stimuli were assigned to rich or lean) and key bindings to rich and lean stimuli were counterbalanced across participants and sessions.

**Figure 1.**
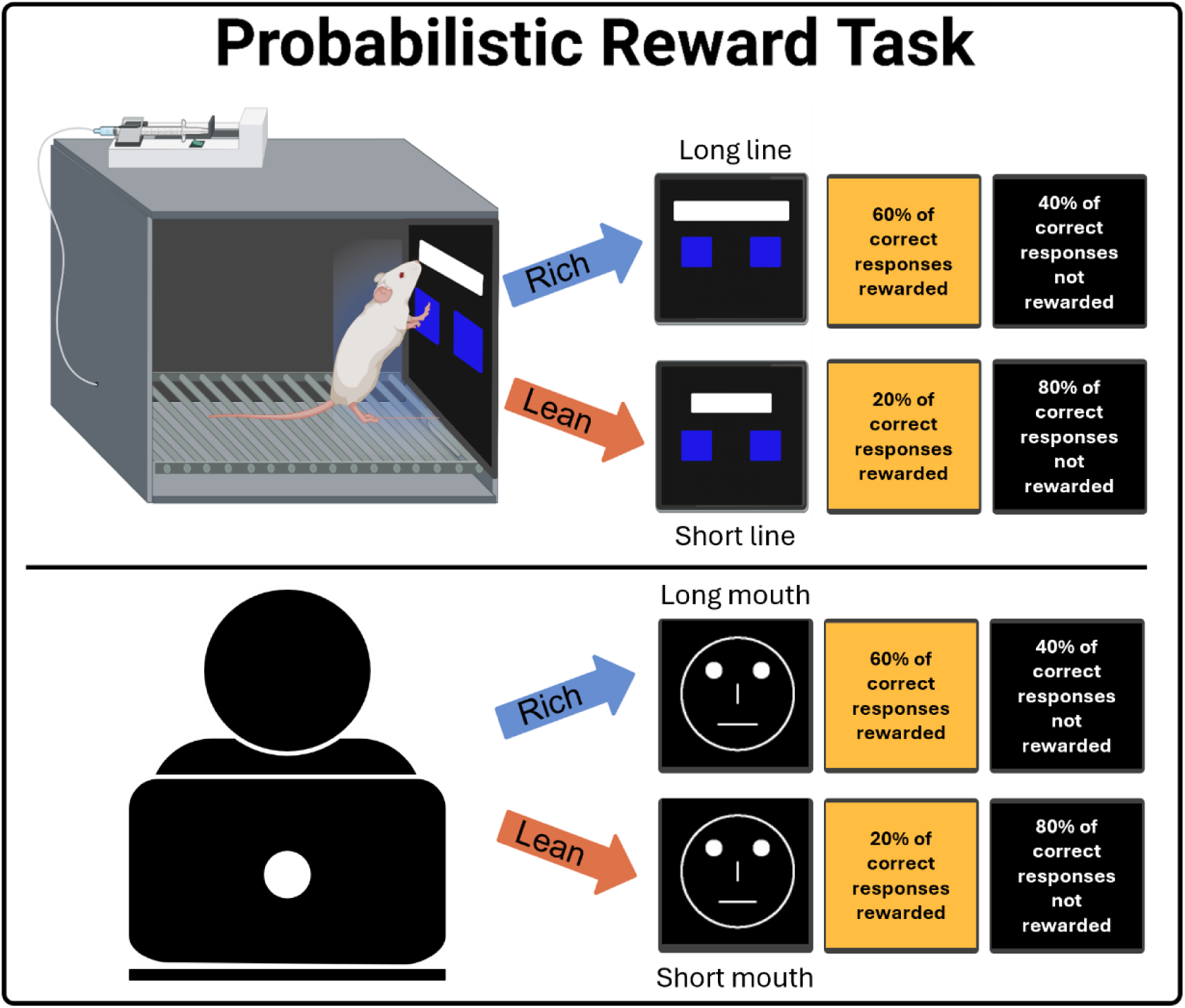
The human and rat version of the Probabilistic Reward Task. Rats responded to short or long line stimuli presented on a touchscreen. Humans judged the length of the mouth stimulus of a schematic face presented on a computer screen. Across species, identical asymmetric reward schedules (3:1) were used to induce a response bias toward the more frequently rewarded stimulus.

For the rat version, subjects completed the PRT in a touch-sensitive experimental chamber (Figure 1) (40,42,46,65). In each session, rats were presented with 100 stimuli that varied in line length (long line: 600×60 px/31.5×3.25 cm; short line: 200×60 px/10.5×3.25 cm. As in the human version, reward contingencies were asymmetric with a 3:1 ratio (rich stimuli: 60% vs. lean stimuli: 20%).

For both PRT versions, the primary measure of interest was response bias, which was computed following classic signal detection theory (43,66,67):

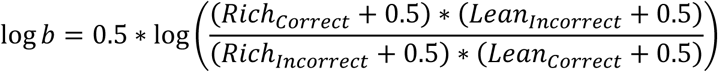

Discriminability, which assesses task difficulty, served as control variable and was computed as:

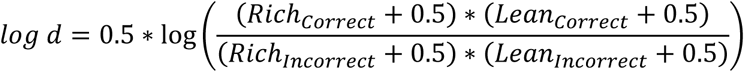

A constant of 0.5 was used to allow log-transformation and avoid division by zero in cases where there are no responses to a given category (68).

### Procedure

Rats were trained in the PRT using previously published protocols (40) (see *Supplement* for more details) until discrimination accuracy reached ≥80% for two consecutive sessions. Animals were then assigned to either testing conditions without programmed stress (n=10, “healthy control” group) or conditions of ongoing chronic stress (n=10, “anhedonic phenotype” group). For the former, PRT testing consisted of three daily 100-trial sessions. For the latter, pre-stress baseline PRT performance was examined during one session before animals were exposed to chronic inescapable ice water stress (48). This involved placing the rat in an opaque polycarbonate cylinder (52 cm high, 40 cm diameter) filled with water iced to 10°C, which is associated with reliable swim durations of approximately 4-8min prior to subsequent submersion. Rats were observed continuously during their swim duration and rescued after submerging for >7s without an apparent ability to resurface. After rescue, animals were placed singly in a clean home cage, without the aid of towel drying or heat lamp, for a 2.5-hour period. This process was repeated daily until a blunted response bias (i.e., at least half of the subject’s pre-stress baseline value) was observed in the PRT. The following day, during continued chronic stress exposure, rats received ketamine at 30min after stress exposure and 2h before behavioral testing. The stress procedure continued the next day, followed by another PRT test session, to examine the effects of ketamine treatment on PRT metrics 24h post-administration.

Human participants completed two testing sessions (2.5h each) separated by 48 hours. For TRD participants, sessions were scheduled 24 hours before and 24 hours after administration of their first dose of ketamine. Healthy participants did not receive any intervention between the two sessions. If the first testing session took place more than one month after screening, participants were re-assessed to confirm continued eligibility. On both days, participants were fitted with a 96-channel electrode cap before they completed 8min of resting state EEG recording, followed by an unrelated flanker task and the PRT. EEG findings will be presented elsewhere. Further, we collected HAMD, QIDS, BDI-II, and SHAPS scores at each testing session.

### Data Analysis

Analyses were performed in R (ver. 4.2.2) using RStudio (ver. 2022.12.0) and the tidyr (69), rstatix (70), lme4 (71) and lmerTest packages (72). Questionnaire and interview scores for human participants were analyzed using mixed-model ANOVAs with group (TRD vs. HC) as between subject factor and session (first vs. second) as within subject factor. Linear mixed effects regression models were used to analyze changes in response bias and discriminability in the PRT across groups and between sessions. Rat and human data were analyzed separately. To align the analysis approach across species, we chose a similar dummy-coding scheme for both data sets. Specifically, we defined the clinical phenotype (i.e., chronically stressed rats and TRD participants) as the baseline category for the *Group* predictor and the first testing session (i.e., pre-stress in rats/pre-treatment in humans) as the baseline category for the *Session* predictor. In this operationalization, effects of *Group* and *Session* in both rats and humans can be similarly interpreted as changes in response bias or discriminability from the respective baseline category, i.e., the “clinical” group at their first testing session (73–75). Note that the number of testing sessions was asymmetric in the rat sample (stressed group: 4 sessions, controls: 3 sessions). Thus, while our models allow assessments of changes in outcomes across all timepoints in the stressed group, there will be no comparison between groups for one of them (here, we chose this timepoint to be the 2h post-injection session in the stress group). For humans, we also included a linear *Block* term in the model (coded as −0.5, 0, 0.5 for blocks 1, 2, and 3, respectively). Regression models included all main and interaction terms as fixed effects as well as a random intercept. Given age group difference in humans, we ran additional regression models controlling for age. Control analyses indicated that age had no significant effect on any outcome measure and did not change the significance level of any of the other predictors in the models (see *Supplement*, Table S3). For simplicity, we thus here report the original models without age.

## Results

### Rats

Regression analyses suggested that, at baseline, rats in the stress group displayed a slightly larger response bias than controls (*Group*: *p*=.049; full results in Table 2, Figure 2A). As expected, response bias in the stress group was significantly reduced following chronic stress exposure (*Session_Stress*: *p*<.001), an effect not seen in unstressed rats (*Group* × *Session_Stress*: *p*<.001). Critically, after ketamine injection, stressed rats’ response bias returned to levels comparable to their baseline (*Session_2_hours* and *Session_24_hours*: *p*=.092 and *p*=.421, respectively). Both groups showed similar degrees of change from their first to their last session (*Group* × *Session_24_hours*: *p*=.505). A targeted post-hoc comparison using a two-sample Welch t-test indicated that response bias in stressed rats was not significantly different from response bias in the third PRT session of the unstressed rats at 24h post-injection (*t*_(11.36)_=2.16, *p*=.053).

**Figure 2.**
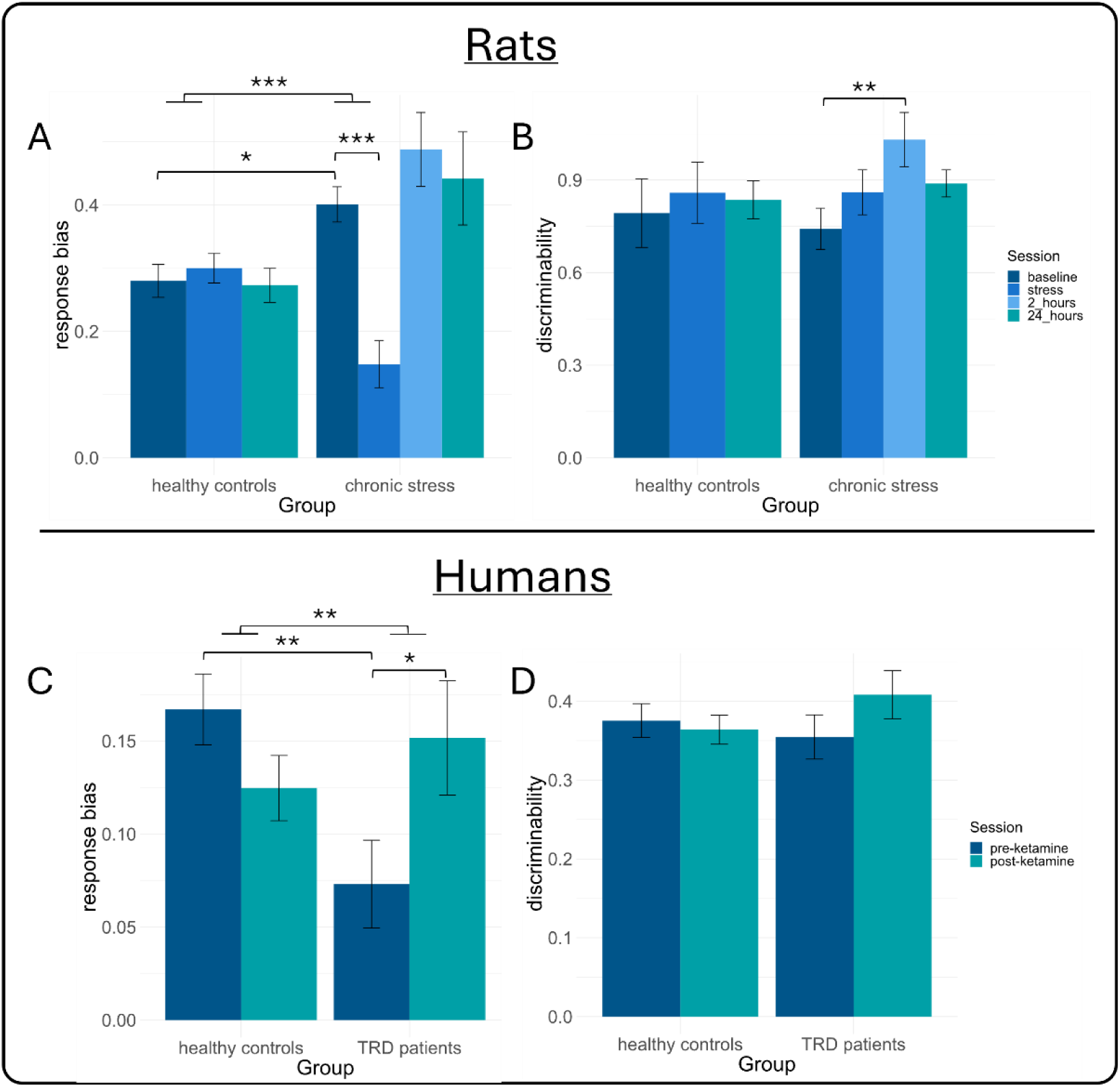
Effects of ketamine administration on response bias and discriminability. In rats, ketamine rescued the stress-induced response bias deficit in the anhedonic phenotype group and returned levels to baseline. The control group displayed stable response bias across three sessions (A). The stress manipulation did not affect discriminability, but it was increased in stressed rats 2h after ketamine administration before returning to baseline (B). In humans, ketamine increased response bias in TRD patients. Response bias in healthy controls did not change significantly between sessions (C). Discriminability remained stable across sessions in both groups (D). * *p*<.05, ** *p*<.01, *** *p*<.001.

**Table 2.** Results from dummy-coded linear mixed model regressions on response bias and discriminability in rats and humans.

| Predictor | *b* (SE) | *df* | *t* | *p* |  |
| --- | --- | --- | --- | --- | --- |
| **Rats** |  |  |  |  |  |
| **Response Bias** |  |  |  |  |  |
| Intercept | 0.401 (0.043) | 52.569 | 9.424 | < .001 | *** |
| Group | -0.121 (0.060) | 52.569 | -2.011 | .049 | * |
| Session_Stress | -0.253 (0.050) | 46.506 | -5.010 | < .001 | *** |
| Session_2_hours | 0.087 (0.050) | 46.506 | 1.723 | .092 |  |
| Session_24_hours | 0.041 (0.050) | 46.506 | 0.812 | .421 |  |
| Group x Session_Stress | 0.273 (0.071) | 46.506 | 3.823 | < .001 | *** |
| Group x Session_24hours | -0.048 (0.071) | 46.506 | -0.672 | .505 |  |
| **Discriminability** |  |  |  |  |  |
| Intercept | 0.742 (0.081) | 40.440 | 9.149 | < .001 | *** |
| Group | 0.051 (0.115) | 40.440 | 0.445 | .659 |  |
| Session_Stress | 0.118 (0.084) | 44.686 | 1.404 | .167 |  |
| Session_2_hours | 0.289 (0.084) | 44.686 | 3.439 | .001 | ** |
| Session_24_hours | 0.147 (0.084) | 44.686 | 1.749 | .087 |  |
| Group x Session_Stress | -0.052 (0.119) | 44.686 | -0.438 | .664 |  |
| Group x Session_24hours | -0.104 (0.119) | 44.686 | -0.875 | .386 |  |
| **Humans** |  |  |  |  |  |
| **Response Bias** |  |  |  |  |  |
| Intercept | 0.073 (0.025) | 145.556 | 2.947 | .004 | ** |
| Group | 0.094 (0.032) | 145.556 | 2.952 | .004 | ** |
| Session_post | 0.079 (0.035) | 223.996 | 2.246 | .026 | * |
| Block | 0.003 (0.030) | 223.996 | 0.123 | .902 |  |
| Group x Session_post | -0.121 (0.045) | 223.996 | -2.697 | .008 | ** |
| Group x Block | 0.019 (0.039) | 223.996 | 0.491 | .624 |  |
| Session_post x Block | -0.004 (0.043) | 223.996 | -0.103 | .918 |  |
| Group x Session_post x Block | 0.012 (0.055) | 223.996 | 0.216 | .829 |  |
| **Discriminability** |  |  |  |  |  |
| Intercept | 0.355 (0.035) | 65.330 | 10.004 | < .001 | *** |
| Group | 0.021 (0.045) | 65.330 | 0.456 | .650 |  |
| Session_post | 0.054 (0.030) | 224.000 | 1.770 | .078 |  |
| Block | -0.002 (0.026) | 224.000 | -0.062 | .951 |  |
| Group x Session_post | -0.065 (0.039) | 224.000 | -1.675 | .095 |  |
| Group x Block | 0.008 (0.034) | 224.000 | 0.238 | .812 |  |
| Session_post x Block | -0.013 (0.037) | 224.000 | -0.357 | .722 |  |
| Group x Session_post x Block | 0.024 (0.047) | 224.000 | 0.495 | .621 |  |
| **Response Bias by SHAPS** |  |  |  |  |  |
| Intercept | 0.077 (0.025) | 136.339 | 3.092 | .002 | ** |
| Group | 0.088 (0.032) | 136.323 | 2.791 | .006 | ** |
| Session_post | 0.088 (0.035) | 232.352 | 2.509 | .013 | * |
| Block | 0.004 (0.030) | 219.936 | 0.125 | .901 |  |
| SHAPS score | -0.003 (0.004) | 137.598 | -0.768 | .444 |  |
| Group x Session_post | -0.126 (0.045) | 228.355 | -2.832 | .005 | ** |
| Group x Block | 0.019 (0.038) | 219.936 | 0.500 | .618 |  |
| Session_post x Block | -0.004 (0.042) | 219.936 | -0.104 | .917 |  |
| Group x SHAPS score | -0.003 (0.005) | 137.278 | -0.553 | .581 |  |
| Session_post x SHAPS score | 0.014 (0.006) | 263.977 | 2.551 | .011 | * |
| Group x Session_post x Block | 0.012 (0.054) | 219.936 | 0.220 | .826 |  |
| Group x Session_post x SHAPS score | -0.013 (0.007) | 260.193 | -1.721 | .086 |  |

**Note.** SHAPS = Snaith-Hamilton Pleasure Scale.

Discriminability did not differ between groups at baseline (*Group*: *p*=.659) and was unaffected by chronic stress exposure (*Session_Stress*: *p*=.167), indicating that stress led to a specific reduction in reward responsiveness in stressed rats but not in their general ability to perform the task (Figure 2B). Ketamine injection enhanced discriminability acutely at 2h post-injection (*Session_2_hours*: *p*=.001), but levels largely returned to baseline at the 24h mark (*Session_24_hours*: *p*=.087). No group effects emerged for discriminability in later sessions (all *p*>.386), suggesting that discriminability remained similarly stable in both groups over time (note that the increased discriminability in stressed rats 2 hours post-injection has no direct comparison in healthy subjects per our modeling choices). We also analyzed the effects of ketamine on subjects’ response times in the PRT (for results, see Table S1, Figure S1).

### Humans

We first examined whether ketamine affected depressive symptom severity scores. Reduction in symptoms were observed for all measures 24h after the first dose (Table 1). Participants with TRD scored higher on all measures compared to HCs (*Group*, HAMD: *F*_(1, 42)_=260.99, *p*<.001, η^2^=0.81; QIDS: *F*_(1, 41)_=306.35, *p*<.001, η^2^=0.81; BDI-II: *F*_(1, 40)_=362.19, *p*<.001, η^2^=0.86; SHAPS: *F*_(1, 44)_=99.16, *p*<.001, η^2^=0.69). Across all participants, we saw reductions in scores for HAMD (*F*_(1, 42)_=18.46, *p*<.001, η^2^=0.13), QIDS (*F*_(1, 41)_=18.81, *p*<.001, η^2^=0.17), and BDI-II (*F*_(1,40)_=7.31, *p*=.010, η^2^=0.06), but not SHAPS (*F*_(1, 44)_=1.77, *p*=.191, η^2^=0.01), indicated by main effects of *Session*. Critically, we found significant *Group* × *Session* interaction effects for all measures, indicating a selective decrease in scores for the TRD participants in session two, but not for HCs (HAMD: *F*_(1, 42)_=16.14, *p*<.001, η^2^=0.11; QIDS: *F*_(1, 41)_=20.05, *p*<.001, η^2^=0.17; BDI-II: *F*_(1, 40)_=7.11, *p*=.011, η^2^=0.05; SHAPS: *F*_(1, 44)_=6.47, *p*=.015, η^2^=0.02).

Our main analysis concerned the effects of ketamine on participants’ response bias (and discriminability; Figure 2C and 2D). As expected, the TRD group displayed a significantly lower response bias compared to HCs in their first session (*Group*: *p*=.004; see Table 2). Twenty-four hours after their first ketamine dose, however, response bias was substantially increased in TRD patients (*Session_post*: *p*=.026). Crucially, this increase was specific to the TRD group (*Group* × *Session*_post: *p*=.008), whereas response bias in healthy controls remained stable between sessions (a targeted post-hoc t-test in HCs revealed that the numerical decrease in response bias between sessions was not significant: (*t*_(45.14)_=-0.35, *p*=.725). We did not find any significant effects involving block (all *ps*>.624).

Discriminability did not differ by group status, session, or block (all *ps*>.078). Overall, these findings mirror the rodent results. Additional results from a regression on RTs are presented in the *Supplement* (Table S2, Figure S2). In brief, TRD participants responded significantly faster in session 2 (*p*=.007). No other effect was significant.

Given the general improvements in patients’ depressive symptom severity between sessions, we explored whether the observed behavioral increase in response bias was modulated by clinical measures. In a set of exploratory analyses, we added participants’ symptom scores to our regression model. These scores were mean-centered across sessions but within groups, given the (intended) large difference in symptom severity between TRD patients and HCs. This allowed proper comparisons of how symptom severity affects response bias in each group (76). We were specifically interested in the SHAPS, given the established link between anhedonia and reduced response bias on the PRT. Findings (Table 2) indicate that SHAPS scores indeed modulated the change in response bias between sessions in the TRD group (*Session_post* × SHAPS score: *p*=.011), suggesting that increases in response bias were larger in more anhedonic patients. This effect was not apparent in the HC group (see Figure 3A and 3B). Notably, separate analyses on each of the other clinical measures (BDI-II, QIDS, HAMD) did not show comparable moderation effects (see *Supplement*, Tables S4-S6), potentially indicating that the post-ketamine increase in the patients’ response bias was specifically related to improvements in anhedonic symptoms.

**Figure 3.**
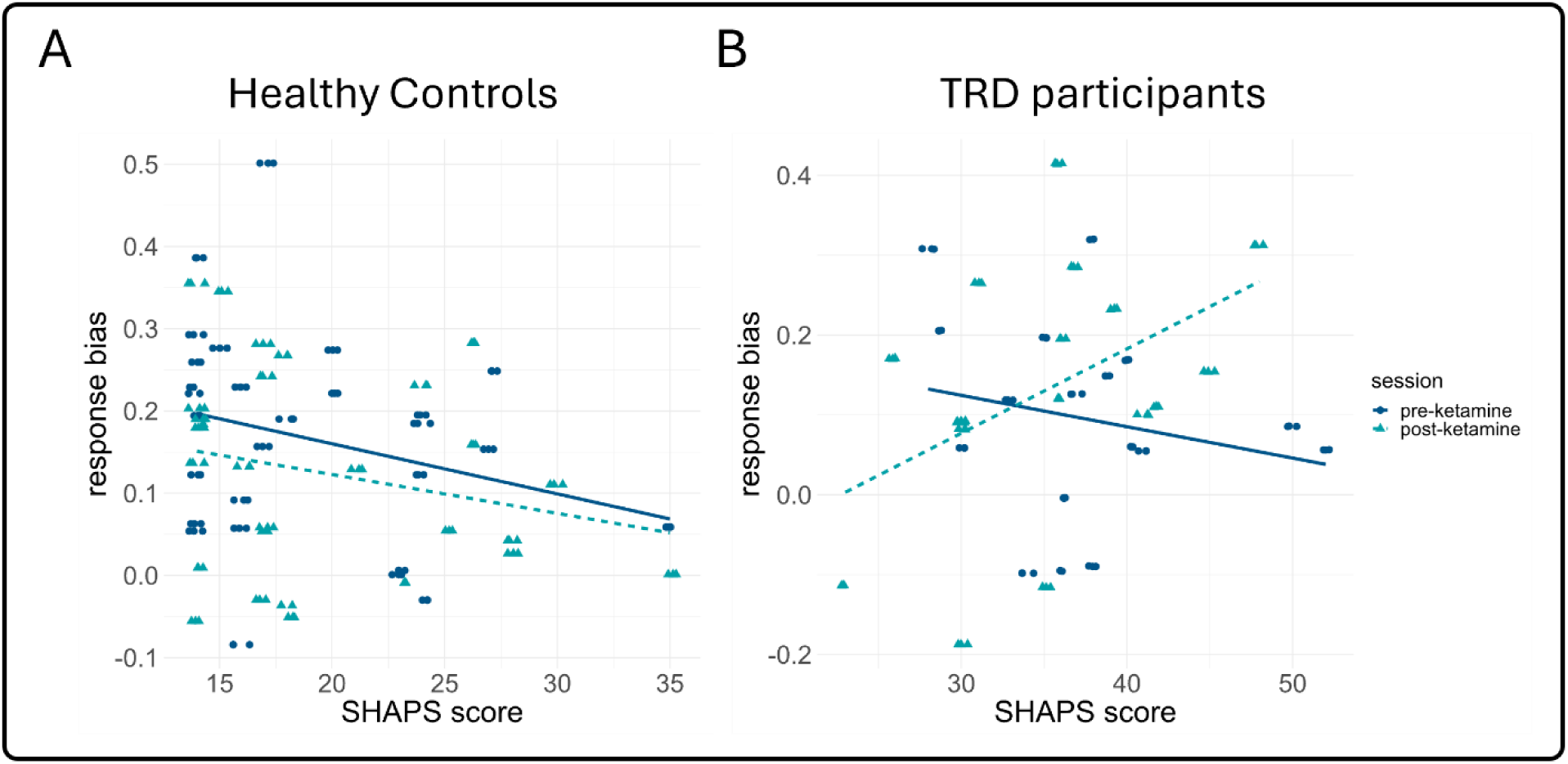
Effects of SHAPS scores on response bias across sessions. For healthy controls, higher SHAPS scores were numerically associated with lower response bias in both sessions (A). In TRD participants, the relationship between SHAPS scores and response bias changed direction following ketamine administration, suggesting highest increases in patients with more severe anhedonia (B).

## Discussion

While ketamine has been shown to exert rapid antidepressant and prohedonic effects in humans and animals (4,5,16,20,20,58,60), the underlying neurocognitive mechanisms are not well understood, partially due to challenges in translating preclinical findings across species (35–37). Here, we investigated whether a single, subanesthetic dose of ketamine would enhance reward responsiveness in people with TRD and rats who displayed anhedonic-like behavior, using functionally identical versions of an established reward paradigm, the PRT (38,40). Consistent with our hypotheses, both species showed a significant increase in response bias toward the rich (i.e., more frequently rewarded) stimulus after ketamine infusion. In rats, this increase was apparent as soon as 2 hours post administration and remained stable after 24 hours, returning response bias magnitude to levels observed prior to chronic stress exposure and comparable to those observed in the control (unstressed) group. Similarly, response bias in TRD participants 24 hours after their first dose of ketamine was comparable to those of healthy controls, equalizing the difference in response bias observed prior to ketamine treatment. Together, our results provide cross-species evidence for a ketamine-induced increase in reward responsiveness, highlighting an evolutionary-conserved mechanism through which ketamine alleviates depressive phenotypes.

In line with previous work, we observed significant reductions in depressive and anhedonic symptoms across all clinical measures following ketamine administration (5,10,12–16,20). However, these measures rely heavily on self-report. In humans, comparatively few studies have investigated the effects of ketamine on more objective markers, such as changes in cognition or behavior that may mediate symptomatic improvements and of those, most have focused on attentional processes or general cognitive capacity rather than reward processing (77–80). In animal models of depression, acute and chronic ketamine administration have been associated with increases in sucrose preference and intake in animals following exposure to chronic stress, implying ketamine might act by modifying responsiveness to anticipated or received reward (15,28,59,81–86). Our results provide evidence that ketamine may act through similar mechanisms in humans, given the near identical patterns of changes in response bias in rats and humans tested with functionally identical versions of the PRT.

Interestingly, ketamine acted specifically on subjects’ reward responsiveness rather than general task difficulty, given that it did not affect discriminability in either species, except for a transient increase in chronically stressed rats at 2h post-administration. Moreover, exploratory analyses revealed that, in TRD participants, ketamine-related changes in response bias were moderated by their SHAPS scores, but not by BDI-II, QIDS, or HAMD, indicating that ketamine improved reward responsiveness primarily in more anhedonic individuals. This is in line with previous work reporting the prohedonic effects of ketamine to be unrelated to its effects on other symptoms of depression (5,26). However, future studies are needed to confirm the specificity of this effect.

On a neural level, response bias in the PRT has been linked to activation in the dorsal ACC and ventral striatum in both humans and rats (36,87). Interestingly, prior work using Positron Emission Tomography in MDD participants and healthy individuals identified ketamine-related modulations of glucose metabolism in various nodes of the reward system, including the dorsal ACC and ventral striatum (6,22,23), highlighting a possible role of these regions in mediating the observed effects of ketamine on response bias. However, we do not propose that the prohedonic properties of ketamine result from specific modulations of isolated brain areas.

Instead, they likely reflect widespread neuroplastic changes within reward networks. Specifically, in rodents, ketamine has been observed to trigger a rapid and transient surge of extracellular glutamate levels and a reversal in stress-related synaptic deficits in the medial prefrontal cortex; moreover, ketamine has been found to decrease burst firing in the lateral habenula, which may drive anhedonic behavior by inhibiting dopaminergic and serotonergic midbrain areas, including the ventral tegmental area and dorsal raphe nuclei (26,28,81,88). Given the evolutionary preserved nature of frontostriatal pathways, it is possible that the observed increases in response bias in both humans and rats resulted from ketamine’s effect on these key reward network nodes that orchestrate activity in downstream areas across the brain. Functional neuroimaging studies will be required to test this conjecture.

Finally, we want to acknowledge some limitations of this study. First, as we relied on referrals of treatment-eligible individuals with TRD instead of active recruitment, the size of our human sample was relatively modest. Further, healthy controls did not receive an intervention comparable to the TRD participants (e.g., saline injection), leaving open the possibility that the observed effects occurred due to procedural differences or placebo effects. Future studies should thus assess larger human samples and implement double-blinded, randomized control designs, ideally including longitudinal measures to test the longevity of acute ketamine administration, given that they have been shown to be short-lived (89,90). Similarly, in the rodent sample, non-stressed rats did not receive ketamine, making it difficult to assess whether the response bias increase observed in stressed rats post-ketamine administration selectively ameliorated stress-induced anhedonia-like behavior or whether ketamine would have induced a more global increase in response bias in both groups. Prior work demonstrated that the dose of ketamine used here (10mg/kg) may engender a response bias increase even in non-stressed rats at 2h, but not 24h post-injection, suggesting partially selective and more persistent improvements in chronically stressed rats (49). Despite these limitations, when taken together, our findings in humans and rats provide converging evidence for similar anti-anhedonic effects of acute ketamine administration in both species. However, to further increase translational utility, future rodent studies on anhedonic-like behavior should include female subjects, given emerging evidence that female rats may be more sensitive to ketamine (91) and the fact that women are disproportionally affected by affective disorders (92,93).

In conclusion, we found translational evidence that ketamine administration rapidly affects reward responsiveness in individuals with TRD and chronically stressed rats. These effects might be specific to ketamine-induced changes in hedonic processes, as we did not see effects on discriminability 24h post-injection and, also, given that the ketamine-related improvements were largest for individuals with more severe anhedonic symptoms. Although more work is necessary to fully characterize the neurobiological, psychological, and computational mechanisms at play, as well as the longevity of these effects, our findings may have important implications for the treatment of those suffering from anhedonia beyond the context of TRD, given that motivational deficits related to anhedonia are commonly observed after stress and across diagnostic domains (94–97). As that there is not yet an approved treatment for these symptoms, our findings may contribute to an objective quantification in the evaluation of novel treatments designed to improve quality of life for people experiencing anhedonia.

## Data Availability Statement

The data used in the present work are available upon reasonable request to the corresponding author.

## Author contributions

DAP and BDK designed the study. JNS, SME, ML, SEW, and ARJ acquired the data. MB, BDK, and DAP analyzed and interpreted the data. MB, BDK, and DAP wrote the initial draft of the manuscript. All authors contributed to the revision and editing of the manuscript and gave final approval before submission.

## References

1. Gaynes BN, Lux L, Gartlehner G, Asher G, Forman-Hoffman V, Green J, et al. (2020): Defining treatment-resistant depression. Depression and Anxiety 37: 134–145.

2. McIntyre RS, Alsuwaidan M, Baune BT, Berk M, Demyttenaere K, Goldberg JF, et al. (2023): Treatment-resistant depression: definition, prevalence, detection, management, and investigational interventions. World psychiatry 22: 394–412.

3. De Carlo V, Calati R, Serretti A (2016): Socio-demographic and clinical predictors of non-response/non-remission in treatment resistant depressed patients: A systematic review. Psychiatry Research 240: 421–430.

4. Berman RM, Cappiello A, Anand A, Oren DA, Heninger GR, Charney DS, Krystal JH (2000): Antidepressant effects of ketamine in depressed patients. Biological Psychiatry 47: 351–354.

5. Lally N, Nugent AC, Luckenbaugh DA, Ameli R, Roiser JP, Zarate CA (2014): Anti-anhedonic effect of ketamine and its neural correlates in treatment-resistant bipolar depression. Transl Psychiatry 4: e469–e469.

6. Lally N, Nugent AC, Luckenbaugh DA, Niciu MJ, Roiser JP, Zarate CA (2015): Neural correlates of change in major depressive disorder anhedonia following open-label ketamine. J Psychopharmacol 29: 596–607.

7. Serafini G, H. Howland R, Rovedi F, Girardi P, Amore M (2014): The Role of Ketamine in Treatment-Resistant Depression: A Systematic Review. Current Neuropharmacology 12: 444–461.

8. Zanos P, Gould TD (2018): Mechanisms of ketamine action as an antidepressant. Mol Psychiatry 23: 801–811.

9. Medeiros GC, Demo I, Goes FS, Zarate CA, Gould TD (2024): Personalized use of ketamine and esketamine for treatment-resistant depression. Transl Psychiatry 14: 1–11.

10. Naughton M, Clarke G, O′Leary OF, Cryan JF, Dinan TG (2014): A review of ketamine in affective disorders: Current evidence of clinical efficacy, limitations of use and pre-clinical evidence on proposed mechanisms of action. Journal of Affective Disorders 156: 24–35.

11. McIntyre RS, Rodrigues NB, Lee Y, Lipsitz O, Subramaniapillai M, Gill H, et al. (2020): The effectiveness of repeated intravenous ketamine on depressive symptoms, suicidal ideation and functional disability in adults with major depressive disorder and bipolar disorder: Results from the Canadian Rapid Treatment Center of Excellence. Journal of Affective Disorders 274: 903–910.

12. Zhuo C, Ji F, Tian H, Wang L, Jia F, Jiang D, et al. (2020): Transient effects of multi-infusion ketamine augmentation on treatment-resistant depressive symptoms in patients with treatment-resistant bipolar depression – An open-label three-week pilot study. Brain and Behavior 10: e01674.

13. Bahji A, Vazquez GH, Zarate CA (2021): Comparative efficacy of racemic ketamine and esketamine for depression: A systematic review and meta-analysis. Journal of Affective Disorders 278: 542–555.

14. Delfino RS, Del-Porto JA, Surjan J, Magalhães E, Sant LCD, Lucchese AC, et al. (2021): Comparative effectiveness of esketamine in the treatment of anhedonia in bipolar and unipolar depression. Journal of Affective Disorders 278: 515–518.

15. Nogo D, Jasrai AK, Kim H, Nasri F, Ceban F, Lui LMW, et al. (2022): The effect of ketamine on anhedonia: improvements in dimensions of anticipatory, consummatory, and motivation-related reward deficits. Psychopharmacology 239: 2011–2039.

16. Patarroyo-Rodriguez L, Cavalcanti S, Vande Voort JL, Singh B (2024): The Use of Ketamine for the Treatment of Anhedonia in Depression. CNS Drugs 38: 583–596.

17. Halahakoon DC, Kieslich K, O’Driscoll C, Nair A, Lewis G, Roiser JP (2020): Reward-Processing Behavior in Depressed Participants Relative to Healthy Volunteers: A Systematic Review and Meta-analysis. JAMA Psychiatry 77: 1286–1295.

18. Guineau MG, Ikani N, Rinck M, Collard R, Van Eijndhoven P, Tendolkar I, et al. (2023): Anhedonia as a transdiagnostic symptom across psychological disorders: a network approach. Psychological medicine 53: 3908–3919.

19. Cao B, Zhu J, Zuckerman H, Rosenblat JD, Brietzke E, Pan Z, et al. (2019): Pharmacological interventions targeting anhedonia in patients with major depressive disorder: A systematic review. Progress in Neuro-Psychopharmacology and Biological Psychiatry 92: 109–117.

20. Rodrigues NB, McIntyre RS, Lipsitz O, Cha DS, Lee Y, Gill H, et al. (2020): Changes in symptoms of anhedonia in adults with major depressive or bipolar disorder receiving IV ketamine: Results from the Canadian Rapid Treatment Center of Excellence. Journal of affective disorders 276: 570–575.

21. Vasavada MM, Loureiro J, Kubicki A, Sahib A, Wade B, Hellemann G, et al. (2021): Effects of serial ketamine infusions on corticolimbic functional connectivity in major depression. Biological Psychiatry: Cognitive Neuroscience and Neuroimaging 6: 735–744.

22. Smith PhD Gwenn S, Schloesser MD Ralf, Brodie PhD M D, Jonathan D, Dewey PhD Stephen L, Logan PhD Jean, Vitkun MD Stephen A, et al. (1998): Glutamate Modulation of Dopamine Measured *in Vivo* with Positron Emission Tomography (PET) and 11C-Raclopride in Normal Human Subjects. Neuropsychopharmacology 18: 18–25.

23. Vollenweider FX, Vontobel P, Øye I, Hell D, Leenders KL (2000): Effects of (S)-ketamine on striatal dopamine: a [11C] raclopride PET study of a model psychosis in humans. Journal of psychiatric research 34: 35–43.

24. Abdallah CG, Averill LA, Collins KA, Geha P, Schwartz J, Averill C, et al. (2017): Ketamine Treatment and Global Brain Connectivity in Major Depression. Neuropsychopharmacol 42: 1210–1219.

25. Abdallah CG, Ahn K-H, Averill LA, Nemati S, Averill CL, Fouda S, et al. (2021): A robust and reproducible connectome fingerprint of ketamine is highly associated with the connectomic signature of antidepressants. Neuropsychopharmacology 46: 478–485.

26. Ballard ED, Wills K, Lally N, Richards EM, Luckenbaugh DA, Walls T, et al. (2017): Anhedonia as a clinical correlate of suicidal thoughts in clinical ketamine trials. Journal of Affective Disorders 218: 195–200.

27. Evans JW, Szczepanik J, Brutsché N, Park LT, Nugent AC, Zarate Jr CA (2018): Default mode connectivity in major depressive disorder measured up to 10 days after ketamine administration. Biological psychiatry 84: 582–590.

28. Yang Y, Cui Y, Sang K, Dong Y, Ni Z, Ma S, Hu H (2018): Ketamine blocks bursting in the lateral habenula to rapidly relieve depression. Nature 554: 317–322.

29. Mkrtchian A, Evans JW, Kraus C, Yuan P, Kadriu B, Nugent AC, et al. (2021): Ketamine modulates fronto-striatal circuitry in depressed and healthy individuals. Molecular psychiatry 26: 3292–3301.

30. Moujaes F, Ji JL, Rahmati M, Burt JB, Schleifer C, Adkinson BD, et al. (2024): Ketamine induces multiple individually distinct whole-brain functional connectivity signatures. elife 13: e84173.

31. Lucantonio F, Roeglin J, Li S, Lu J, Shi A, Czerpaniak K, et al. (2025): Ketamine rescues anhedonia by cell-type- and input-specific adaptations in the nucleus accumbens. Neuron 0. 10.1016/j.neuron.2025.02.021

32. Duncan Jr WC, Slonena E, Hejazi NS, Brutsche N, Yu KC, Park L, et al. (2017): Motor-activity markers of circadian timekeeping are related to ketamine’s rapid antidepressant properties. Biological Psychiatry 82: 361–369.

33. Can A, Zanos P, Moaddel R, Kang HJ, Dossou KS, Wainer IW, et al. (2016): Effects of ketamine and ketamine metabolites on evoked striatal dopamine release, dopamine receptors, and monoamine transporters. The Journal of Pharmacology and Experimental Therapeutics 359: 159–170.

34. Xu S, Yao X, Li B, Cui R, Zhu C, Wang Y, Yang W (2022): Uncovering the underlying mechanisms of ketamine as a novel antidepressant. Frontiers in Pharmacology 12: 740996.

35. Pizzagalli DA (2022): Toward a Better Understanding of the Mechanisms and Pathophysiology of Anhedonia: Are We Ready for Translation? AJP 179: 458–469.

36. Iturra-Mena AM, Kangas BD, Luc OT, Potter D, Pizzagalli DA (2023): Electrophysiological signatures of reward learning in the rodent touchscreen-based Probabilistic Reward Task. Neuropsychopharmacol 48: 700–709.

37. Rizvi SJ, Pizzagalli DA, Sproule BA, Kennedy SH (2016): Assessing anhedonia in depression: Potentials and pitfalls. Neuroscience & Biobehavioral Reviews 65: 21–35.

38. Pizzagalli DA, Jahn AL, O’Shea JP (2005): Toward an objective characterization of an anhedonic phenotype: A signal-detection approach. Biological Psychiatry 57: 319–327.

39. Insel TR (2014): The NIMH Research Domain Criteria (RDoC) Project: Precision Medicine for Psychiatry. AJP 171: 395–397.

40. Kangas BD, Wooldridge LM, Luc OT, Bergman J, Pizzagalli DA (2020): Empirical validation of a touchscreen probabilistic reward task in rats. Translational Psychiatry 10: 285.

41. Wooldridge LM, Bergman J, Pizzagalli DA, Kangas BD (2021): Translational Assessments of Reward Responsiveness in the Marmoset. International Journal of Neuropsychopharmacology 24: 409–418.

42. Luc OT, Kangas BD (2024): Validation of a touchscreen probabilistic reward task for mice: A reverse-translated assay with cross-species continuity. *Cognitive, Affective*, & Behavioral Neuroscience 24: 281–288.

43. Luc OT, Pizzagalli DA, Kangas BD (2021): Toward a quantification of anhedonia: unified matching law and signal detection for clinical assessment and drug development. Perspectives on Behavior Science 44: 517–540.

44. Pizzagalli DA, Iosifescu D, Hallett LA, Ratner KG, Fava M (2008): Reduced hedonic capacity in major depressive disorder: evidence from a probabilistic reward task. Journal of psychiatric research 43: 76–87.

45. Vrieze E, Pizzagalli DA, Demyttenaere K, Hompes T, Sienaert P, de Boer P, et al. (2013): Reduced Reward Learning Predicts Outcome in Major Depressive Disorder. Biological Psychiatry 73: 639–645.

46. Kangas BD, Short AK, Luc OT, Stern HS, Baram TZ, Pizzagalli DA (2022): A cross-species assay demonstrates that reward responsiveness is enduringly impacted by adverse, unpredictable early-life experiences. Neuropsychopharmacology 47: 767–775.

47. Lamontagne SJ, Wash SI, Irwin SH, Zucconi KE, Olmstead MC (2022): Effects of dopamine modulation on chronic stress-induced deficits in reward learning. *Cognitive, Affective*, & Behavioral Neuroscience 22: 736–753.

48. Gonzalez PM, Jenkins AR, LaMalfa KS, Kangas BD (2024): Chronic ecologically relevant stress effects on reverse-translated touchscreen assays of reward responsivity and attentional processes in male rats: Implications for depression. Journal of Neurochemistry 168: 2190–2200.

49. Jenkins AR, Radl DB, Kornecook TJ, Pizzagalli DA, Bergman J, Buhl DL, et al. (2025): Environmental determinants of ketamine’s prohedonic and antianhedonic efficacy: Persistence of enhanced reward responsiveness is modulated by chronic stress. The Journal of Pharmacology and Experimental Therapeutics 392: 103572.

50. National Research Council, Division on Earth, Life Studies, Institute for Laboratory Animal Research (2011): Guidance for the description of animal research in scientific publications.

51. Sheehan DV, Lecrubier Y, Sheehan KH, Amorim P, Janavs J, Weiller E, et al. (1998): The Mini-International Neuropsychiatric Interview (MINI): the development and validation of a structured diagnostic psychiatric interview for DSM-IV and ICD-10. Journal of clinical psychiatry 59: 22–33.

52. Hamilton M (1960): A rating scale for depression. Journal of neurology, neurosurgery, and psychiatry 23: 56.

53. Beck AT, Steer RA, Brown GK (1996): Manual for the beck depression inventory-II.

54. Rush AJ, Trivedi MH, Ibrahim HM, Carmody TJ, Arnow B, Klein DN, et al. (2003): The 16-Item quick inventory of depressive symptomatology (QIDS), clinician rating (QIDS-C), and self-report (QIDS-SR): a psychometric evaluation in patients with chronic major depression. Biological Psychiatry 54: 573–583.

55. Snaith R, Hamilton M, Morley S, Humayan A, Hargreaves D, Trigwell P (1995): A scale for the assessment of hedonic tone the Snaith–Hamilton Pleasure Scale. The British Journal of Psychiatry 167: 99–103.

56. Pizzagalli DA, Smoski M, Ang Y-S, Whitton AE, Sanacora G, Mathew SJ, et al. (2020): Selective kappa-opioid antagonism ameliorates anhedonic behavior: evidence from the Fast-fail Trial in Mood and Anxiety Spectrum Disorders (FAST-MAS). Neuropsychopharmacology 45: 1656–1663.

57. Dillon DG, Lazarov A, Dolan S, Bar-Haim Y, Pizzagalli DA, Schneier FR (2022): Fast evidence accumulation in social anxiety disorder enhances decision making in a probabilistic reward task. Emotion 22: 1.

58. Garcia LS, Comim CM, Valvassori SS, Réus GZ, Barbosa LM, Andreazza AC, et al. (2008): Acute administration of ketamine induces antidepressant-like effects in the forced swimming test and increases BDNF levels in the rat hippocampus. Progress in neuro-psychopharmacology and biological psychiatry 32: 140–144.

59. Garcia LS, Comim CM, Valvassori SS, Réus GZ, Stertz L, Kapczinski F, et al. (2009): Ketamine treatment reverses behavioral and physiological alterations induced by chronic mild stress in rats. Progress in Neuro-Psychopharmacology and Biological Psychiatry 33: 450–455.

60. Wang J, Goffer Y, Xu D, Tukey DS, Shamir D, Eberle SE, et al. (2011): A single sub-anesthetic dose of ketamine relieves depression-like behaviors induced by neuropathic pain in rats. Anesthesiology 115: 812.

61. Fava M, Freeman MP, Flynn M, Judge H, Hoeppner BB, Cusin C, et al. (2020): Double-blind, placebo-controlled, dose-ranging trial of intravenous ketamine as adjunctive therapy in treatment-resistant depression (TRD). Mol Psychiatry 25: 1592–1603.

62. Sanacora G, Frye MA, McDonald W, Mathew SJ, Turner MS, Schatzberg AF, et al. (2017): A consensus statement on the use of ketamine in the treatment of mood disorders. JAMA psychiatry 74: 399–405.

63. Su T-P, Chen M-H, Li C-T, Lin W-C, Hong C-J, Gueorguieva R, et al. (2017): Dose-related effects of adjunctive ketamine in Taiwanese patients with treatment-resistant depression. Neuropsychopharmacology 42: 2482–2492.

64. Wilkinson ST, Farmer C, Ballard ED, Mathew SJ, Grunebaum MF, Murrough JW, et al. (2019): Impact of midazolam vs. saline on effect size estimates in controlled trials of ketamine as a rapid-acting antidepressant. Neuropsychopharmacology 44: 1233–1238.

65. Kangas BD, Bergman J (2017): Touchscreen technology in the study of cognition-related behavior. Behavioural pharmacology 28: 623–629.

66. McCarthy D, Davison M (1979): Signal probability, reinforcement and signal detection. Journal of the experimental analysis of behavior 32: 373–386.

67. McCarthy D (1983): Measures of response bias at minimum-detectable luminance levels in the pigeon. Journal of the Experimental Analysis of Behavior 39: 87–106.

68. Hautus MJ (1995): Corrections for extreme proportions and their biasing effects on estimated values ofd′. Behavior Research Methods, Instruments, & Computers 27: 46–51.

69. Wickham H, Vaughan D, Girlich M (2020): tidyr: Tidy Messy Data. R package version 1.1. 3. CRAN R-project org/package= tidyr.

70. Kassambara A (2019): rstatix: Pipe-friendly framework for basic statistical tests. CRAN: Contributed Packages.

71. Bates D, Maechler M (2009): Package ‘lme4’(Version 0.999375-32): linear mixed-effects models using S4 classes. *Available (April 2011) at* http://cran *r-project org/web/packages/lme4/lme4 pdf*.

72. Kuznetsova A, Brockhoff PB, Christensen RH (2017): lmerTest package: tests in linear mixed effects models. Journal of statistical software 82: 1–26.

73. Gelman A, Hill J (2006): Data Analysis Using Regression and Multilevel/Hierarchical Models. Cambridge university press.

74. Sharp ME, Foerde K, Daw ND, Shohamy D (2015): Dopamine selectively remediates ‘model-based’reward learning: a computational approach. Brain 139: 355–364.

75. Bogdanov M, LoParco S, Otto AR, Sharp M (2022): Dopaminergic medication increases motivation to exert cognitive control by reducing subjective effort costs in Parkinson’s patients. Neurobiology of Learning and Memory 193: 107652.

76. Chen G, Adleman NE, Saad ZS, Leibenluft E, Cox RW (2014): Applications of multivariate modeling to neuroimaging group analysis: a comprehensive alternative to univariate general linear model. Neuroimage 99: 571–588.

77. Shiroma PR, Albott CS, Johns B, Thuras P, Wels J, Lim KO (2014): Neurocognitive performance and serial intravenous subanesthetic ketamine in treatment-resistant depression. International Journal of Neuropsychopharmacology 17: 1805–1813.

78. Chen M-H, Li C-T, Lin W-C, Hong C-J, Tu P-C, Bai Y-M, et al. (2018): Cognitive function of patients with treatment-resistant depression after a single low dose of ketamine infusion. Journal of Affective Disorders 241: 1–7.

79. Gill H, Gill B, Rodrigues NB, Lipsitz O, Rosenblat JD, El-Halabi S, et al. (2021): The Effects of Ketamine on Cognition in Treatment-Resistant Depression: A Systematic Review and Priority Avenues for Future Research. Neuroscience & Biobehavioral Reviews 120: 78–85.

80. Singh B, Parikh SV, Voort JLV, Pazdernik VK, Achtyes ED, Goes FS, et al. (2024): Change in neurocognitive functioning in patients with treatment-resistant depression with serial intravenous ketamine infusions: The Bio-K multicenter trial. Psychiatry Research 335: 115829.

81. Li N, Liu R-J, Dwyer JM, Banasr M, Lee B, Son H, et al. (2011): Glutamate N-methyl-D-aspartate receptor antagonists rapidly reverse behavioral and synaptic deficits caused by chronic stress exposure. Biological psychiatry 69: 754–761.

82. Sarkar A, Kabbaj M (2016): Sex differences in effects of ketamine on behavior, spine density, and synaptic proteins in socially isolated rats. Biological psychiatry 80: 448–456.

83. Papp M, Gruca P, Lason-Tyburkiewicz M, Willner P (2017): Antidepressant, anxiolytic and procognitive effects of subacute and chronic ketamine in the chronic mild stress model of depression. Behavioural Pharmacology 28: 1–8.

84. Tornese P, Sala N, Bonini D, Bonifacino T, La Via L, Milanese M, et al. (2019): Chronic mild stress induces anhedonic behavior and changes in glutamate release, BDNF trafficking and dendrite morphology only in stress vulnerable rats. The rapid restorative action of ketamine. Neurobiology of stress 10: 100160.

85. Aricioğlu F, Yalcinkaya C, Ozkartal CS, Tuzun E, Sirvanci S, Kucukali CI, Utkan T (2020): NLRP1-mediated antidepressant effect of ketamine in chronic unpredictable mild stress model in rats. Psychiatry investigation 17: 283.

86. Dalla Vecchia D, Kanazawa LKS, Wendler E, Hocayen P de AS, Vital MABF, Takahashi RN, et al. (2021): Ketamine reversed short-term memory impairment and depressive-like behavior in animal model of Parkinson’s disease. Brain Research Bulletin 168: 63–73.

87. Santesso DL, Evins AE, Frank MJ, Schetter EC, Bogdan R, Pizzagalli DA (2009): Single dose of a dopamine agonist impairs reinforcement learning in humans: Evidence from event-related potentials and computational modeling of striatal-cortical function. Human Brain Mapping 30: 1963–1976.

88. Li N, Lee B, Liu R-J, Banasr M, Dwyer JM, Iwata M, et al. (2010): mTOR-dependent synapse formation underlies the rapid antidepressant effects of NMDA antagonists. Science 329: 959–964.

89. Zarate CA, Singh JB, Carlson PJ, Brutsche NE, Ameli R, Luckenbaugh DA, et al. (2006): A randomized trial of an N-methyl-D-aspartate antagonist in treatment-resistant major depression. Archives of general psychiatry 63: 856–864.

90. Ibrahim L, DiazGranados N, Franco-Chaves J, Brutsche N, Henter ID, Kronstein P, et al. (2012): Course of improvement in depressive symptoms to a single intravenous infusion of ketamine vs add-on riluzole: results from a 4-week, double-blind, placebo-controlled study. Neuropsychopharmacology 37: 1526–1533.

91. Ponton E, Turecki G, Nagy C (2022): Sex Differences in the Behavioral, Molecular, and Structural Effects of Ketamine Treatment in Depression. International Journal of Neuropsychopharmacology 25: 75–84.

92. Piccinelli M, Wilkinson G (2000): Gender differences in depression: Critical review. The British Journal of Psychiatry 177: 486–492.

93. Hyde JS, Mezulis AH (2020): Gender differences in depression: biological, affective, cognitive, and sociocultural factors. Harvard review of psychiatry 28: 4–13.

94. Shafiei N, Gray M, Viau V, Floresco SB (2012): Acute stress induces selective alterations in cost/benefit decision-making. Neuropsychopharmacology 37: 2194–2209.

95. Husain M, Roiser JP (2018): Neuroscience of apathy and anhedonia: a transdiagnostic approach. Nature Reviews Neuroscience 19: 470.

96. Bogdanov M, Nitschke JP, LoParco S, Bartz JA, Otto AR (2021): Acute psychosocial stress increases cognitive-effort avoidance. Psychological Science 32: 1463–1475.

97. Kuhn M, Palermo EH, Pagnier G, Blank J M, Steinberger DC, Long Y, et al. (2025): Computational Phenotyping of Effort-Based Decision Making in Unmedicated Adults with Remitted Depression. Biological Psychiatry: Cognitive Neuroscience and Neuroimaging. 10.1016/j.bpsc.2025.02.006

