## Supplemental material for "Ketamine improves anhedonic phenotypes across species: Translational evidence from the Probabilistic Reward Task"

[#] Equal senior contributions

### Supplemental Methods

Group-level and individual exclusion criteria for human participants

For screening purposes, exclusion criteria for both groups consisted of: current substance use or lifetime history of seizures, psychosis, bipolar disorder, or unstable medical illness. Healthy controls were excluded if they reported of ≥ 7 (i.e., a cut-off for minimal depression) in the Hamilton Depressive Rating Scale (HAMD*(1)*), a history of psychiatric illness, or currently use of psychotropic medication. TRD participants were required to meet criteria for a current major depressive episode and to have not experienced a 50% reduction in depressive symptoms following at least two full trials of any antidepressant in the current episode, as assessed by the MGH Antidepressant Treatment Response Questionnaire (ATRQ *(2)*).

For analysis purposes, participants were excluded if, in any of the two experimental sessions, they had less than 80% valid trials in a given block of the PRT, received reward feedback in less than 20 rich or 6 lean trials per block, or if their reward ratio of rich vs. lean trials fell below 2:1 in any block.

Sample size was determined based on the requirements of the parent study and is comparable to previous experimental work on ketamine in TRD (3–6) and is sufficient to detect differences in PRT performance between clinical and healthy individuals(7,8).

Training procedure in the rat sample

Using previously published protocols (9), rats were first trained to respond on the touchscreen and, subsequently, to discriminate line stimuli. Trial types varied in a quasi-random manner across 100-trial sessions, with 50 trials of each type. Subjects were differentially reinforced to respond to the left or right response box depending on the length of the white line. Correct responses yielded small amounts of a palatable food reward (0.1 mL of a 30% sweetened condensed milk solution) that was paired with an 880 ms yellow screen flash and 440 Hz tone and followed by a 5-sec blackout period, whereas each incorrect response immediately resulted in a 10-sec blackout period. Training continued until discrimination accuracy was ≥80% for 2 consecutive sessions (7,10,11).

### Supplemental Results

In addition to response bias and discriminability, we also performed regression analyses on response time (RT) in the PRT. For this purpose, we added trial type (rich vs. lean, effect-coded as 0.5 and -0.5, respectively) as a predictor to our regression models, which were otherwise identical to the ones used for response bias and discriminability.

Rats

For the rat sample, we used median RT as the dependent variable (see also Figure S1). Results suggest that, at baseline, rats in the stress group responded faster to rich compared to lean stimuli (main effect Trial Type: $p$ = .014), which can be seen as a further manifestation of a preference in favor of the more frequently rewarded stimulus. This RT advantage for rich trials was observed across both groups and sessions as indicated by the absence of significant interaction effects involving trial type (i.e., indicating that the difference in RTs seen in the baseline category did not change by group or session, all $p$ > .206). We also found a significant Group x Session_24hours interaction ($p$ = .019), indicating an increase in RTs for both rich and lean stimuli in unstressed rats compared to a decrease in stressed rats, in relation to their respective baseline session. We did not observe any changes in RTs in stressed rats following ketamine administration (main effects Session_2_hours and Session_24_hours: p > .214)

**Table S1.** Results from a linear mixed-effects regression predicting median RT by Group, Session, and Trial Type in the rat sample.

| Predictor | *b (SE)* | *df* | *t* | *p* | |
|---|---|---|---|---|---|
| Intercept | 1.409 (0.252) | 37.398 | 5.597 | < .001 | *** |
| Group | 0.377 (0.356) | 37.398 | 1.059 | .296 | |
| Session_Stress | -0.161 (0.238) | 108.092 | -0.675 | .501 | |
| Session_2_hours | -0.298 (0.238) | 108.092 | -1.250 | .214 | |
| Session_24_hours | -0.251 (0.238) | 108.092 | -1.055 | .294 | |
| Trial Type | -0.838 (0.337) | 108.092 | -2.490 | .014 | * |
| Group x Session_Stress | 0.222 (0.337) | 108.092 | 0.660 | .511 | |
| Group x Session_24hours | 0.801 (0.337) | 108.092 | 2.380 | .019 | * |
| Group x Trial Type | 0.478 (0.476) | 108.092 | 1.004 | .317 | |
| Session_Stress x Trial Type | 0.605 (0.476) | 108.092 | 1.271 | .206 | |
| Session_2_hours x Trial Type | 0.403 (0.476) | 108.092 | 0.847 | .399 | |
| Session_24_hours x Trial Type | 0.596 (0.476) | 108.092 | 1.252 | .213 | |
| Group x Session_Stress x Trial Type | -0.278 (0.673) | 108.092 | -0.413 | .680 | |
| Group x Session_24hours x Trial Type | -0.278 (0.673) | 108.092 | -0.413 | .680 | |

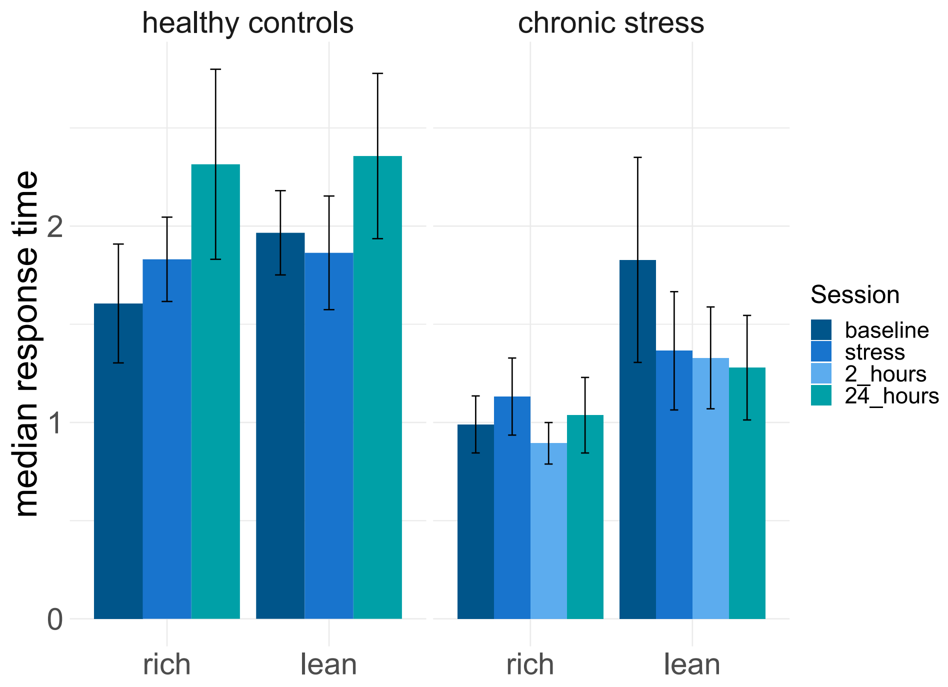


**Figure S1.** Median response times for each group of rats and session split by trial type. Note that in controls, there was no equivalent to the 2h post-ketamine session in the stressed rats.

<u>Humans</u>

In humans, we used natural-log-transformed RTs as the dependent variable (see also Figure S2). To reduce model complexity, and because we did not find effects of block in the initial analysis, we excluded block from the model. Results suggested TRD participants responded overall faster in their second session (i.e., after ketamine administration), independent of trial type (main effect Session_post: $p$ = .007). No other effects were significant (all $p$s > .248).

**Table S2.** Results from a linear mixed-effects regression predicting lnRT by Group, Session, and Trial Type in the human sample.

| **Predictor** | ***b (SE)*** | ***df*** | ***t*** | ***p*** | |
|---|---|---|---|---|---|
| Intercept | 6.324 (0.047) | 47.780 | 134.181 | < .001 | *** |
| Group | 0.012 (0.060) | 47.780 | 0.194 | .847 | |
| Session_post | -0.053 (0.019) | 132.000 | -2.765 | .007 | ** |
| Trial type | -0.031 (0.027) | 132.000 | -1.160 | .248 | |
| Group x Session_post | 0.018 (0.024) | 132.000 | 0.733 | .465 | |
| Group x Trial type | 0.005 (0.034) | 132.000 | 0.143 | .887 | |
| Session_post x Trial type | 0.010 (0.038) | 132.000 | 0.256 | .798 | |
| Group x Session_post x Trial type | -0.005 (0.049) | 132.000 | -0.103 | .918 | |

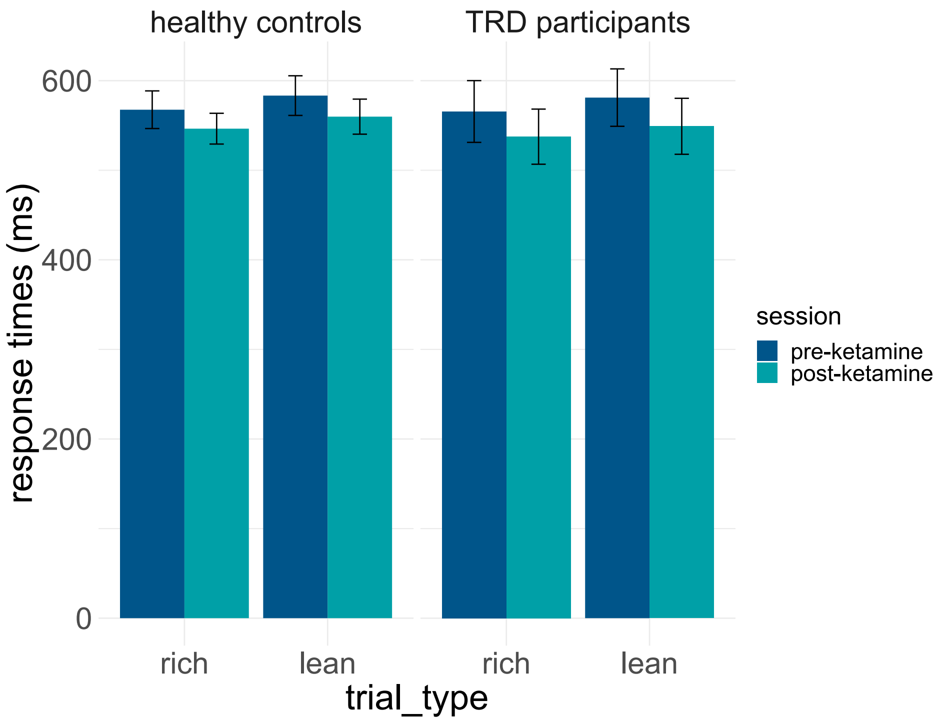


**Figure S2.** Response times for each group of participants and session split by trial type.

Age effects

Given the significant difference in age between groups, we performed control analyses that additionally included age at assessment (mean-centered across all participants) as a main effect and in interaction with group and session to investigate whether it changes our results. We did not find any significant effects of age in these analyses (Table S3).

**Table S3.** Results from linear mixed-effects regression models controlling for age effects on response bias and discriminability.

| | Predictor | b (SE) | df | t | p | |
|---|---|---|---|---|---|---|
| **Response Bias** | | | | | | |
| | Intercept | 0.073 (0.025) | 134.172 | 2.926 | .004 | ** |
| | Group | 0.094 (0.032) | 134.172 | 2.931 | .004 | ** |
| | Session | 0.079 (0.035) | 218.000 | 2.263 | .025 | * |
| | Block | 0.004 (0.030) | 218.000 | 0.124 | .902 | |
| | Age | 0.000 (0.002) | 134.172 | 0.007 | .995 | |
| | Group x session | -0.121 (0.044) | 218.000 | -2.716 | .007 | ** |
| | Group x block | 0.019 (0.039) | 218.000 | 0.495 | .621 | |
| | Session x block | -0.004 (0.042) | 218.000 | -0.103 | .918 | |
| | Group x age | -0.003 (0.002) | 134.172 | -1.258 | .211 | |
| | Session x age | 0.001 (0.002) | 218.000 | 0.458 | .647 | |
| | Block x age | 0.002 (0.002) | 218.000 | 1.310 | .192 | |
| | Group x session x block | 0.012 (0.054) | 218.000 | 0.218 | .828 | |
| | Group x session x age | 0.004 (0.003) | 218.000 | 1.469 | .143 | |
| | Group x block x age | -0.002 (0.003) | 218.000 | -0.723 | .471 | |
| | Session x block x age | -0.003 (0.003) | 218.000 | -1.055 | .292 | |
| | Group x session x block x age | 0.002 (0.004) | 218.000 | 0.698 | .486 | |
| **Discriminability** | | | | | | |
| | Intercept | 0.355 (0.036) | 62.267 | 9.981 | < .001 | *** |
| | Group | 0.021 (0.046) | 62.267 | 0.455 | .651 | |
| | Session | 0.054 (0.030) | 218.000 | 1.770 | .078 | |
| | Block | -0.002 (0.026) | 218.000 | -0.062 | .951 | |
| | Age | -0.002 (0.002) | 62.267 | -0.905 | .369 | |
| | Group x session | -0.065 (0.039) | 218.000 | -1.674 | .096 | |
| | Group x block | 0.008 (0.034) | 218.000 | 0.238 | .812 | |
| | Session x block | -0.013 (0.037) | 218.000 | -0.357 | .722 | |
| | Group x age | 0.003 (0.003) | 62.267 | 1.022 | .311 | |
| | Session x age | -0.001 (0.002) | 218.000 | -0.546 | .586 | |
| | Block x age | -0.001 (0.002) | 218.000 | -0.269 | .789 | |
| | Group x session x block | 0.024 (0.048) | 218.000 | 0.495 | .621 | |
| | Group x session x age | -0.003 (0.003) | 218.000 | -1.037 | .301 | |
| | Group x block x age | 0.001 (0.002) | 218.000 | 0.274 | .785 | |
| | Session x block x age | 0.002 (0.002) | 218.000 | 0.841 | .401 | |
| | Group x session x block x age | -0.003 (0.003) | 218.000 | -0.844 | .400 | |

Additional clinical measures

As with Snaith-Hamilton Pleasure Scale (SHAPS; (12), we included the remaining clinical measures (Hamilton Depressive Rating Scale (HAMD(1)), Beck Depression Inventory (BDI(13)), Quick-Inventory of Depression (QIDS(14))) in the regression models to evaluate whether changes in response bias were associated with depressive symptoms. However, we did not find any modulations related to those measures (tables S5 – S6).

**Table S4.** Response bias by BDI

| Predictor | *b (SE)* | *df* | *t* | *p* | |
|---|---|---|---|---|---|
| Intercept | 0.057 (0.027) | 252.000 | 2.100 | .037 | * |
| Group | 0.106 (0.034) | 252.000 | 3.149 | .002 | ** |
| Session_post | 0.116 (0.037) | 252.000 | 3.116 | .002 | ** |
| Block | 0.003 (0.031) | 252.000 | 0.096 | .924 | |
| BDI score | 0.003 (0.003) | 252.000 | 0.935 | .351 | |
| Group x Session_post | -0.153 (0.047) | 252.000 | -3.251 | .001 | ** |
| Group x Block | 0.019 (0.040) | 252.000 | 0.482 | .630 | |
| Session_post x Block | -0.004 (0.043) | 252.000 | -0.084 | .933 | |
| Group x BDI score | -0.005 (0.018) | 252.000 | -0.280 | .779 | |
| Session_post x BDI score | 0.005 (0.004) | 252.000 | 1.186 | .237 | |
| Group x Session_post x Block | 0.013 (0.056) | 252.000 | 0.231 | .817 | |
| Group x Session_post x BDI score | 0.004 (0.026) | 252.000 | 0.172 | .864 | |

**Note.** BDI = Beck Depression Inventory.

**Table S5.** Response bias by QIDS

| Predictor | *b (SE)* | *df* | *t* | *p* | |
|---|---|---|---|---|---|
| Intercept | 0.100 (0.030) | 255.000 | 3.274 | .001 | ** |
| Group | 0.060 (0.037) | 255.000 | 1.647 | .100 | |
| Session_post | 0.055 (0.042) | 255.000 | 1.298 | .194 | |
| Block | -0.004 (0.031) | 255.000 | -0.118 | .906 | |
| QIDS score | -0.012 (0.007) | 255.000 | -1.669 | .095 | |
| Group x Session_post | -0.091 (0.051) | 255.000 | -1.788 | .074 | |
| Group x Block | 0.025 (0.040) | 255.000 | 0.635 | .526 | |
| Session_post x Block | -0.001 (0.044) | 255.000 | -0.023 | .982 | |
| Group x QIDS score | -0.049 (0.046) | 255.000 | -1.055 | .291 | |
| Session_post x QIDS score | 0.014 (0.009) | 255.000 | 1.527 | .127 | |
| Group x Session_post x Block | 0.009 (0.057) | 255.000 | 0.168 | .867 | |
| Group x Session_post x QIDS score | 0.072 (0.056) | 255.000 | 1.275 | .202 | |

**Note.** QIDS = Quick-Inventory of Depression.

Table **S6.** Response bias by HAMD

| Predictor | *b (SE)* | *df* | *t* | *p* | |
|---|---|---|---|---|---|
| Intercept | 0.087 (0.029) | 128.801 | 2.992 | .003 | ** |
| Group | 0.077 (0.036) | 127.750 | 2.164 | .032 | * |
| Session_post | 0.052 (0.040) | 257.117 | 1.298 | .196 | |
| Block | 0.004 (0.030) | 215.168 | 0.123 | .903 | |
| HAMD score | -0.006 (0.006) | 143.093 | -0.981 | .328 | |
| Group x Session_post | -0.090 (0.049) | 250.773 | -1.826 | .069 | |
| Group x Block | 0.018 (0.039) | 215.168 | 0.461 | .645 | |
| Session_post x Block | -0.008 (0.044) | 215.168 | -0.194 | .846 | |
| Group x HAMD score | -0.017 (0.028) | 134.794 | -0.622 | .535 | |
| Session_post x HAMD score | 0.001 (0.008) | 216.078 | 0.180 | .857 | |
| Group x Session_post x Block | 0.017 (0.056) | 215.168 | 0.304 | .761 | |
| Group x Session_post x HAMD score | 0.038 (0.043) | 257.015 | 0.872 | .384 | |

**Note.** HAMD = Hamilton Depression Rating Scale.

#### References


1. Hamilton M (1960): A rating scale for depression. *Journal of Neurology, Neurosurgery, and Psychiatry* 23: 56.
2. Chandler GM, Iosifescu DV, Pollack MH, Targum SD, Fava M (2010): Validation of the Massachusetts General Hospital Antidepressant Treatment History Questionnaire (ATRQ). *CNS Neuroscience & Therapeutics* 16: 322–325.
3. Murrough JW, Wan L-B, Iacoviello B, Collins KA, Solon C, Glicksberg B, *et al.* (2014): Neurocognitive effects of ketamine in treatment-resistant major depression: association with antidepressant response. *Psychopharmacology* 231: 481–488.
4. Shiroma PR, Albott CS, Johns B, Thuras P, Wels J, Lim KO (2014): Neurocognitive performance and serial intravenous subanesthetic ketamine in treatment-resistant depression. *International Journal of Neuropsychopharmacology* 17: 1805–1813.
5. Chen M-H, Li C-T, Lin W-C, Hong C-J, Tu P-C, Bai Y-M, *et al.* (2018): Cognitive function of patients with treatment-resistant depression after a single low dose of ketamine infusion. *Journal of Affective Disorders* 241: 1–7.
6. Wilkinson ST, Farmer C, Ballard ED, Mathew SJ, Grunebaum MF, Murrough JW, *et al.* (2019): Impact of midazolam vs. saline on effect size estimates in controlled trials of ketamine as a rapid-acting antidepressant. *Neuropsychopharmacology* 44: 1233–1238.
7. Pizzagalli DA, Iosifescu D, Hallett LA, Ratner KG, Fava M (2008): Reduced hedonic capacity in major depressive disorder: evidence from a probabilistic reward task. *Journal of Psychiatric Research* 43: 76–87.
8. Montemitro C, Ossola P, Ross TJ, Huys QJ, Fedota JR, Salmeron BJ, *et al.* (2024): Longitudinal changes in reinforcement learning during smoking cessation: A computational analysis using a probabilistic reward task. *Scientific Reports* 14: 32171.
9. Kangas BD, Wooldridge LM, Luc OT, Bergman J, Pizzagalli DA (2020): Empirical validation of a touchscreen probabilistic reward task in rats. *Translational Psychiatry* 10: 285.

10. Pizzagalli DA, Jahn AL, O'Shea JP (2005): Toward an objective characterization of an anhedonic phenotype: A signal-detection approach. *Biological Psychiatry* 57: 319–327.

11. Pizzagalli DA, Smoski M, Ang Y-S, Whitton AE, Sanacora G, Mathew SJ, *et al.* (2020): Selective kappa-opioid antagonism ameliorates anhedonic behavior: evidence from the Fast-fail Trial in Mood and Anxiety Spectrum Disorders (FAST-MAS). *Neuropsychopharmacology* 45: 1656–1663.

12. Snaith R, Hamilton M, Morley S, Humayan A, Hargreaves D, Trigwell P (1995): A scale for the assessment of hedonic tone the Snaith–Hamilton Pleasure Scale. *The British Journal of Psychiatry* 167: 99–103.

13. Beck AT, Steer RA, Brown GK (1996): Manual for the beck depression inventory-II.

14. Rush AJ, Trivedi MH, Ibrahim HM, Carmody TJ, Arnow B, Klein DN, *et al.* (2003): The 16-Item quick inventory of depressive symptomatology (QIDS), clinician rating (QIDS-C), and self-report (QIDS-SR): a psychometric evaluation in patients with chronic major depression. *Biological Psychiatry* 54: 573–583.
